# Humoral immune response after COVID-19 infection or BNT162b2 vaccine among older adults: evolution over time and protective thresholds

**DOI:** 10.1101/2021.11.19.21266252

**Authors:** Maxence Meyer, Florentin Constancias, Claudia Worth, Anita Meyer, Marion Muller, Alexandre Boussuge, Georges Kaltenbach, Elise Schmitt, Saïd Chayer, Aurélie Velay, Thomas Vogel, Samira Fafi-Kremer, Patrick Karcher

## Abstract

**INTRODUCTION:** The objectives of this study were to assess the dynamics of the SARS-CoV-2 anti-RBD IgG response over time among older people after COVID-19 infection or vaccination and its comparison with speculative levels of protection assumed by current data.

**METHODS:** From November 2020 to October 2021, we included geriatric patients with serological test results for COVID-19.

We considered antibody titre thresholds thought to be high enough to protect against SARS-CoV-2 infection: 141 BAU/ml for protection/vaccine efficacy > 89.3%.

Three cohorts are presented. A vaccine group (n=34) that received two BNT162b2/Comirnaty injections 21 days apart, a group of natural COVID-19 infection (n=32) and a third group who contracted COVID-19 less than 15 days after the first BNT162b2/Comirnaty injection (n=17).

**RESULTS:** 83 patients were included, the median age was 87 (81-91) years.

In the vaccine group at 1 month since the first vaccination, the median BAU/ml with IQR was 620 (217-1874) with 87% of patients above the threshold of 141 BAU/ml. Seven months after the first vaccination the BAU/ml was 30 (19-58) with 9.5% of patients above the threshold of 141 BAU/ml.

In the natural COVID-19 infection group, at 1 month since the date of first symptom onset, the median BAU/ml was 798 (325-1320) with 86.7% of patients above the threshold of 141 BAU/ml and fell to 88 (37-385) with 42.9% of patients above the threshold of 141 BAU/ml at 2 months. The natural infection group was vaccinated three months after the infection. Five months after the end of the vaccination cycle the BAU/ml was 2048 (471-4386) with 83.3% of patients above the threshold of 141 BAU/ml.

**DISCUSSION:** On the humoral level, this supports the clinical results describing the decrease in vaccine protection over time.

## INTRODUCTION

A better description of the kinetics of the anti-SARS-CoV-2 humoral response over time and its correlation with potential protection against COVID-19 among older people is necessary. SARS-CoV-2 anti-RBD (Receptor Binding Domain of the spike protein) IgG are correlated with neutralizing antibodies^1,2^ and are thought to be correlated with protection against COVID-19.^2,3^

The objectives of this study were to assess the dynamics of the SARS-CoV-2 anti-RBD IgG response over time among older people after COVID-19 infection or vaccination and its comparison with speculative levels of protection assumed by current data^2,3^.

## METHODS

The observational cohort study was approved by the institutional ethics board of the University Hospital of Strasbourg. Consent was collected for all patients.

From November 2020 to October 2021, we included all geriatric hospital patients with serological test results for COVID-19 and a history of COVID-19 or COVID vaccination. Serum samples were tested using the SARS-CoV-2 IgG II Quant assay, detecting IgG antibodies directed against the spike RBD of SARS-CoV-2 with results expressed in Binding Antibody Units/ml (BAU/ml) as recommended by the WHO^4^. This allows interlaboratory comparison

Based on data from previous studies, we considered 3 antibody titre thresholds thought to be high enough to protect against SARS-CoV-2 infection: 141 BAU/ml for protection/vaccine efficacy > 89.3% as suggested in the study by Dimeglio et al.^3^ and 165 BAU/ml and 506 BAU/ml, respectively, for a protection/vaccine efficacy of 70% and 80% according to Feng et al.^2^

## RESULTS

83 patients were included, including 60 women (72%). The median age and Charlson Comorbidity Index with Interquartile Range (IQR) was 87 (81-91) years and 7 (5-8.5).

### Serological assessment and antibody titers

The results in the different groups as a function of time are shown in Figure 1 and Table S1.

**Figure 1.**
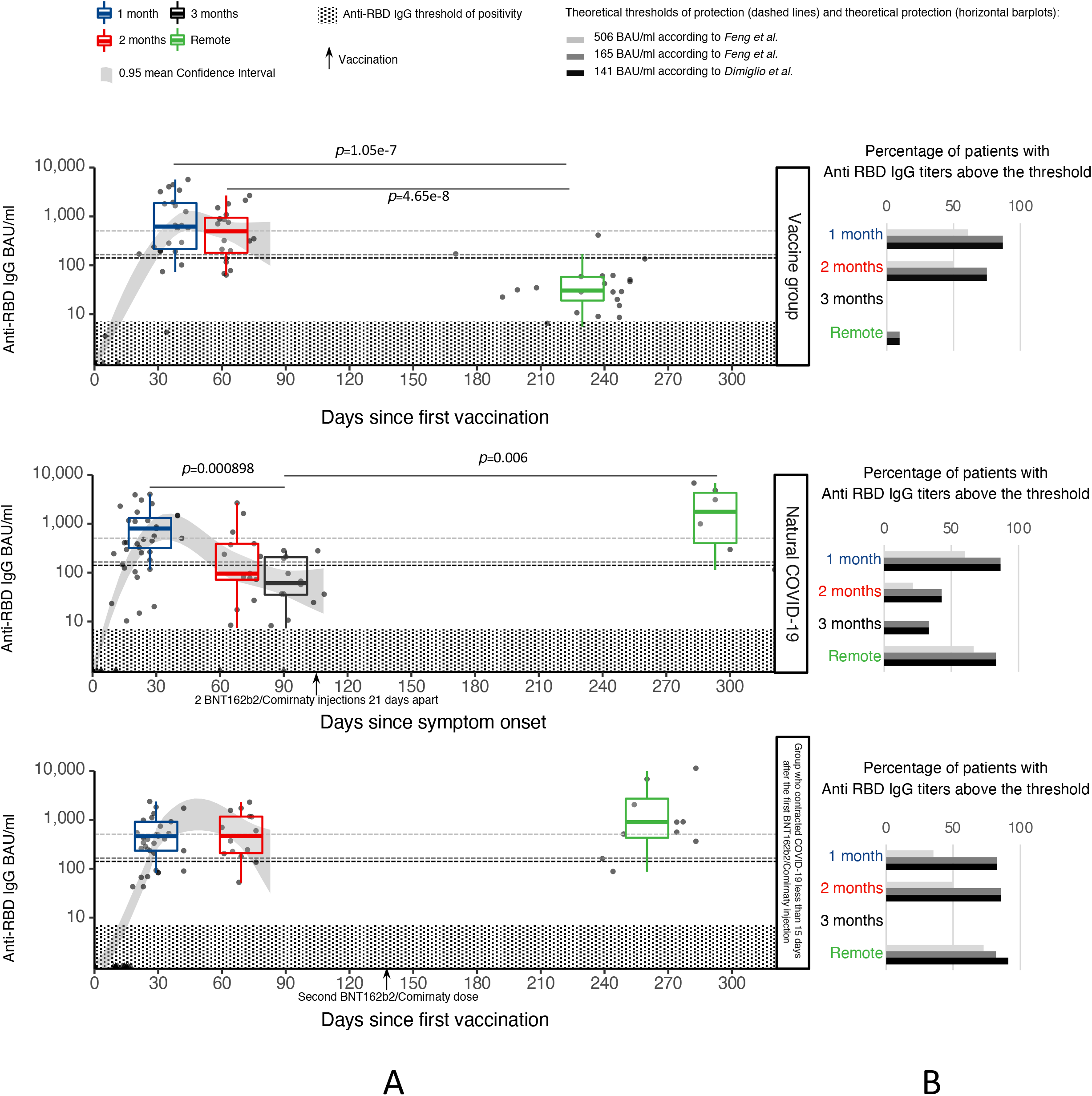
Kinetics of anti-SARS-CoV-2 RBD IgG and theoretical protection against COVID-19 among older adults Panel A: Evolution of humoral response (Anti-RBD IgG BAU/ml) over time. Time axis reflects days since symptom onset (Natural COVID) or after BNT162b2 vaccine first injection (Vaccine group and Group who contracted COVID-19 less than 15 days after the first BNT162b2/Comirnaty injection). Arrows in the x axis represent 2 BNT162b2/Comirnaty injections 21 days apart (Natural COVID) or the second BNT162b2/Comirnaty injection (Group who contracted COVID-19 less than 15 days after the first BNT162b2/Comirnaty injection) Boxplots display Anti-RBD IgG BAU distribution at 1, 2, 3 months or during the post period (first quartile, median, third quartile and whiskers represent 1.5 times the interquartile range from the first and third quartiles). Confidence interval of the mean (0.95 level) was estimated for each group using loess method to describe the dynamics of the humoral response during the first months. Significant differences between periods were estimated within each group using pairwise non-parametric Kruskall-Wallis tests. Holm-adjusted p values indicate significant differences. Horizontal dashed lines represent theoretical threshold of protection against COVID-19 as suggested by *Dimiglio et al*. (141 BAU/ml for protection/vaccine efficacy > 89.3%) and by *Feng et al*. (165 BAU/ml and 506 BAU/ml for protection/vaccine efficacy of 70% and 80%). Panel B: Percentage of patients with theoretical protection against COVID-19=percentage of patients with Anti-RBD IgG titers above a protective threshold for each group, periods and different theoretical thresholds as defined in panel A.

In the vaccine group at 1 month since the first vaccination, the median BAU/ml with IQR was 620 (217-1874) with 87% of patients above the threshold of 141 BAU/ml. At 2 months since the first vaccination the BAU/ml was 526 (182-945) with 75% of patients above the threshold of 141 BAU/ml. Seven months after the first vaccination the BAU/ml was 30 (19-58) with 9.5% of patients above the threshold of 141 BAU/ml.

In the natural COVID-19 infection group, at 1 month since the date of first symptom onset, the median BAU/ml was 798 (325-1320) with 86.7% of patients above the threshold of 141 BAU/ml and fell to 88 (37-385) with 42.9% of patients above the threshold of 141 BAU/ml at 2 months. At three months the BAU/ml was 56 (29-203) with 33.3% of patients above the threshold of 141 BAU/ml. The natural infection group was vaccinated three months after the infection. Five months after the end of the vaccination cycle the BAU/ml was 2048 (471-4386) with 83.3% of patients above the threshold of 141 BAU/ml.

In the group who contracted COVID-19 less than 15 days after the first BNT162b2/Comirnaty injection, at 1 month from the first injection, the median BAU/ml was 463 (234-914) with 82.4% of patients above the threshold of 141 BAU/ml. At two months the BAU/ml was 484 (208-1167) with 85.7% of patients above the threshold of 141 BAU/ml. They received their second dose at three months. 4.5 months after the second dose the BAU/ml was 898 (437-2824) with 90.9% of patients above the threshold of 141 BAU/ml.

## DISCUSSION

Seven months after the first vaccination in COVID-naive patients, the RBD antibody titer decreases with a median BAU/ml of 30 and only 9.5% of patients above the theoretical protective threshold of 141 BAU/ml. A drop by a factor of 20 compared to the peak values wich is consistent with the literature^5^. In COVID-19 patients, a median 56 BAU/ml is observed at three months with 33.3% of patients above the threshold of 141 BAU/ml. However COVID-19 patients, vaccinated at a distance from their infections present, five months post completion of the the vaccination schedule, antibody levels higher than peak levels observed at one month in those exposed to natural infection or standard vaccination alone.

On the humoral level, this supports the clinical results describing the decrease in vaccine protection over time^6^.

### Limitations

Protective thresholds are currently very speculative.

Also even if the BAUs and the thresholds were to become clearer, these thresholds could change over time as the variants evolve and the humoral avidity could decrease in addition to the indisputable humoral decrease.

Sample size is limited.

## Data Availability

All data produced in the present study are available upon reasonable request to the authors

## Conflict of Interest

The authors do not have any conflicts of interest.

## Author contributions

All authors made substantial contributions to this article

The sponsor was Strasbourg University Hospital. The sponsor had no role on the design, methods, subject recruitment, data collections, analysis and preparation of the paper.

## Supplementary Table legends

**Supplementary Table S1.**
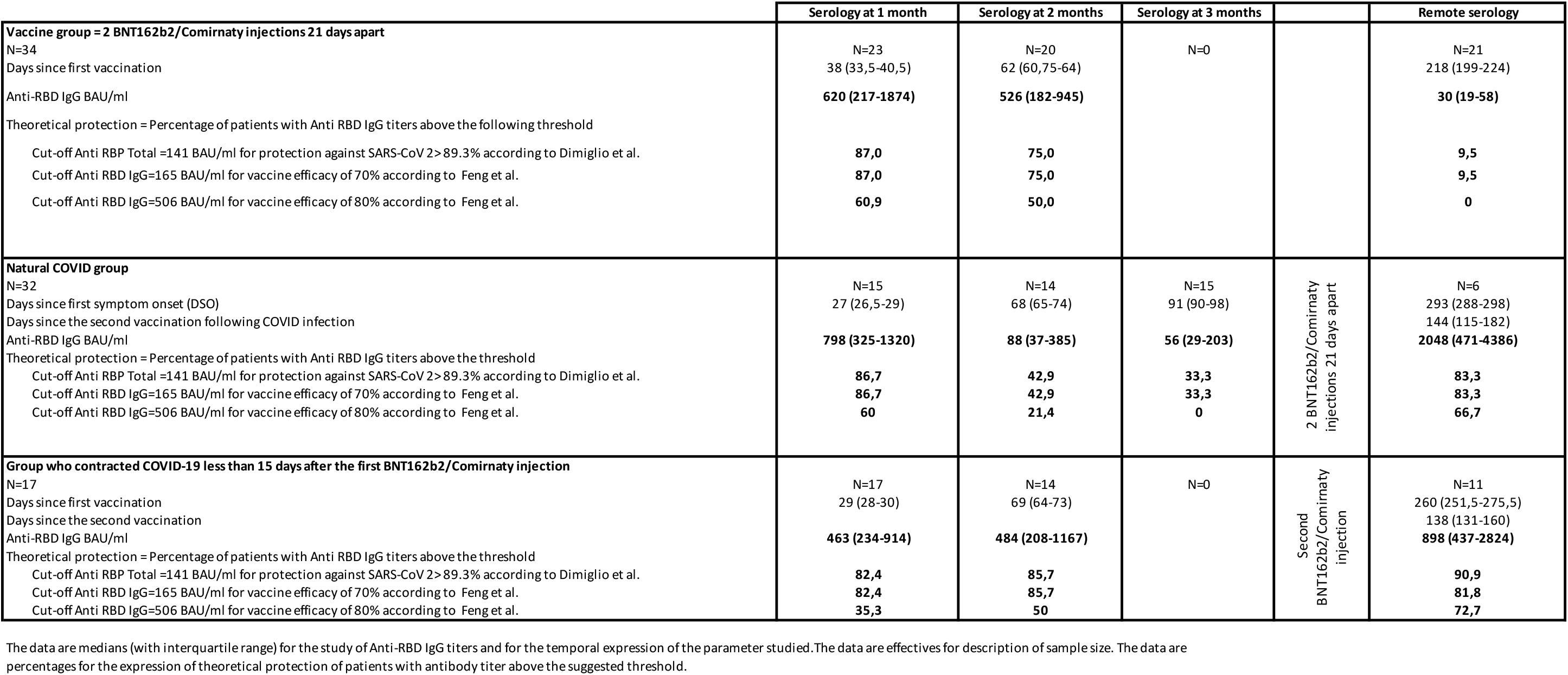
Antibody titers and theoretical protection against COVID-19 among older adults

## Notes

### Competing Interest Statement

The authors have declared no competing interest.

### Funding Statement

This study did not receive any funding

### Author Declarations

Ethics committee of the University Hospital of Strasbourg gave ethique approval for this work

